# Prediction of COVID-19 Mortality to Support Patient Prognosis and Triage and Limits of Current Open-Source Data

**DOI:** 10.1101/2021.03.21.21253984

**Authors:** Riccardo Doyle

## Abstract

This study examines the accuracy and applicability of machine learning methods in early prediction of mortality in COVID-19 patients. Patient symptoms, pre-existing conditions, age and sex were employed as predictive attributes from data spanning 17 countries. Performance on a semi-evenly balanced class sample of 212 patients resulted in high detection accuracy of 92.5%, with strong specificity and sensitivity. Performance on a larger sample of 5,121 patients with only age and mortality information was added as a measure of baseline discriminatory ability. Stratifying - Random Forest - and linear - Logistic Regression - methods were applied, both achieving modestly strong performance, with 77.4%-79.3% sensitivity and 71.4%-72.6% accuracy, highlighting predictive power even on the basis of a single attribute. Mutual information was employed as a dimensionality reduction technique, greatly improving performance and showing how a small number of easily retrievable attributes can provide timely and accurate predictions, with applications for datasets with slowly available variables - such as laboratory results.

Unlike existing studies making use of the same dataset, limitations of the data were extensively explored and detailed, as each results section outlines the main shortcomings of relevant analysis. Future use of this dataset should be cautious and always accompanied by disclaimers on issues of real-life reproducibility. While its open-source nature is a credit to the wider research community and more such datasets should be published, in its current state it can produce valid conclusions only for a limited set of applications, some of which were explored in this study.

## 1 Introduction

The continuing development of the COVID-19 pandemic has tested the limits of hospital resources and staff across the world, placing large importance on effective prognosis for triage and management of admitted patients.

A large number of mortality predictive models that rely on easily available diagnostic and demographic information have been proposed to address the issue, but to varying degrees of usability. Many such models suffer from mild to severe flaws including the lack of patient level variables, training on pneumonia as a proxy for COVID-19, (Barda et. al, 2020), depending on less immediately available data from blood tests and other monitoring equipment (Knight et. al, 2020), unrepresentative – often older skewing (El-Solh, 2020) or mono-localized (An, 2020) – population samples, resulting either in low performance or, more worryingly, in excessively optimistic expectations of performance that overfit to a certain facet of the population.

While all predictive models will inevitably suffer from issues surrounding quality of data or population reproducibility, many of these still generate valuable findings that can materially aid in patient profiling and optimization of treatment and have been adopted on a supportive level by hospitals.

In this study, we aim to further the performance of predictive modelling by addressing the main shortcomings of previous iterations, namely lack of dimensionality reduction pre-processing and data limitations such as lack of geographic diversity, representative age range and other baseline characteristics, leading to more robust performance on unseen data and real-life application. At the same time, we will outline limitations in the data of this study and of a wider number of studies that rely on the same source but make scarce mention of flaws or representativeness issues.

## 2 Methods

### 2.1 Dataset

Data for this study was obtained from a continuously updated repository (Xu, 2020) containing anonymized patient level information on 2,676,311 COVID-19 positive individuals across 146 countries.

### 2.2 Variable Extraction and Data Pre-Processing

Symptoms and comorbidities in the dataset were parsed and one-hot encoded into fixed variable names. The table (Table 1) below shows all patient variables used in the study:

**Table 1:**
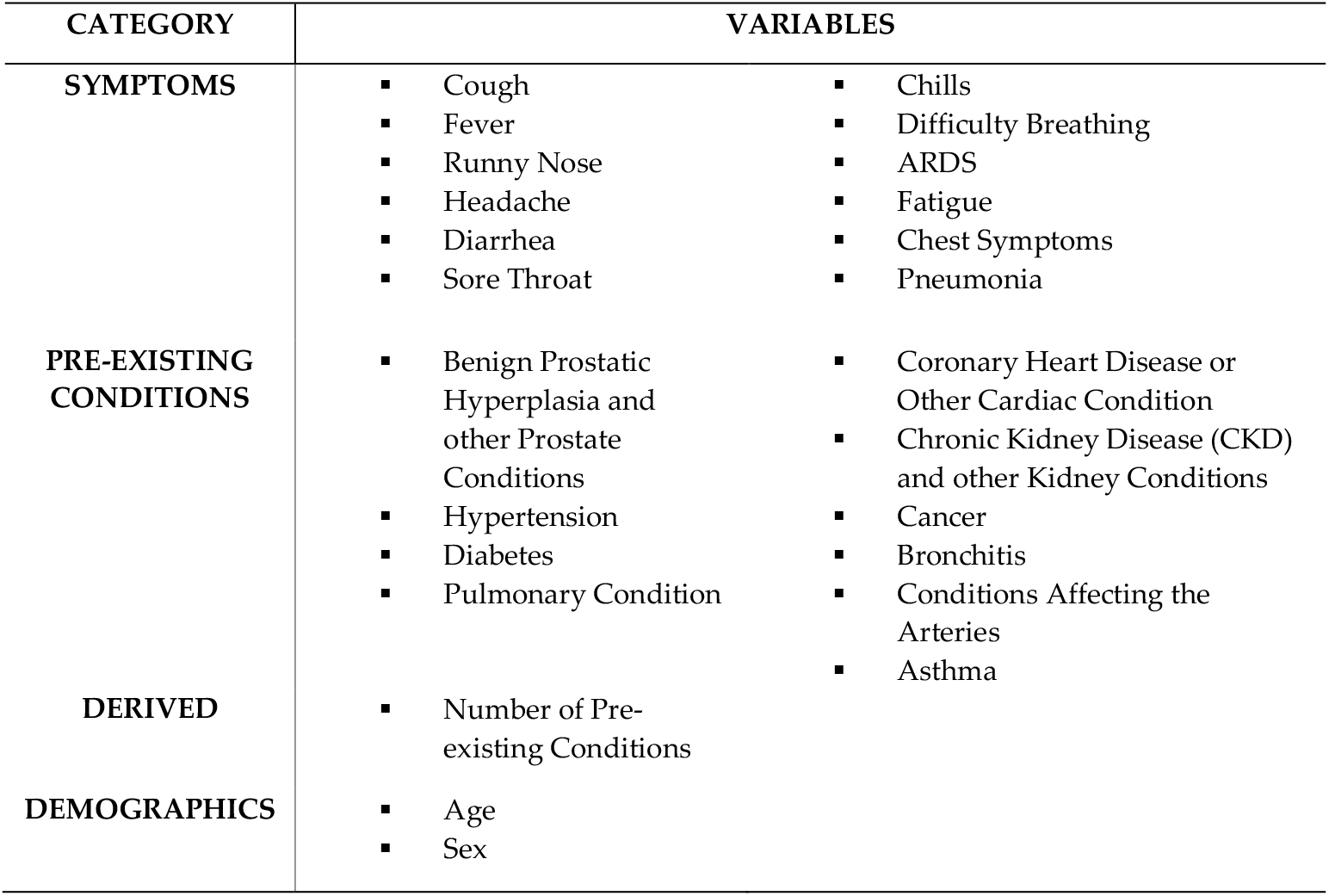
Attributes used as predictors of mortality by category.

As part of our initial analysis, only patients with full data for each of the outlined variables were kept in the study. Of the original 2,676,311 patients, 212 patients satisfied the data quality requirement and were retained in the study. The data is evenly balanced in its proportion of recoveries and deaths, which is not representative.

Finally, a third data subset is used, which includes patients with information on all data categories from Table 1, excluding symptom data. This results in a larger sample of 5,121 patients. A full breakdown of sample characteristics is provided in later results sections.

### 2.3 Dimensionality Reduction

Excessive numbers of explanatory variables can compromise model performance, so Mutual Information was applied to reduce the original 25 patient variables.

The use of variable reduction in this study is partly aimed at avoiding the curse of dimensionality, but primarily at exploring how a smaller number of variables may still carry strong predictive power. Specifically to models that involve laboratory tests and mid to long term variables, the ability to rely on patient history or a smaller number of more immediate variables could expedite the decision making process or act as a first risk screening while results are expected. Two dimensionalities will be compared, one with no reduction and one with 7 features.

### 2.4 Predictive Models

Random Forest and Logistic Regression will be employed as classifiers for this study, the former for its proven performance in the relevant literature, and the latter owing to its statistical nature and to provide a linear counterpart. All classifiers will be trained on ex-ante balanced data and tested on unprocessed imbalanced data.

### 2.5 Evaluation Criteria

Model performance will be evaluated on several key metrics. As the dataset is imbalanced and recovery outcomes far outweigh death outcomes, accuracy is not a reliable measure of performance; a naïve recovery predictor would achieve strong accuracy without providing any benefit.

Sensitivity and specificity will be the focus of model performance. Sensitivity measures the proportion of deaths correctly identified by the model, expressed as:

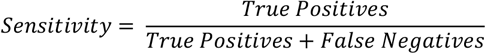

Where death is a positive outcome and recovery is a negative outcome. Specificity measures the proportion of recoveries correctly identified, expressed as:

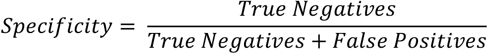

Receiver Operator Characteristic curves will be plotted for some classifiers and the area under the curve (AUC) will serve as an additional point of comparison.

All above metrics will be derived from aggregation during 3-fold to 5-fold Cross Validation depending on size of each dataset variation mentioned earlier.

## 3 Results

### 3.1 Correlation Matrix of Patient Characteristics

Before analysing prediction model performance, the figure below (Fig. 1) outlines the main cross correlation of patient characteristics and their correlation with an outcome of death. We note that the most explanatory features that raise mortality risk are age (correlation coefficient of 0.51), whether a patient has a pre-existing chronic condition (0.59) and the number of pre-existing conditions (0.53). This is followed by particularly risk elevating conditions such as diabetes and hypertension and specific symptoms of advanced disease progression such as pneumonia and ARDS.

**Fig. 1:**
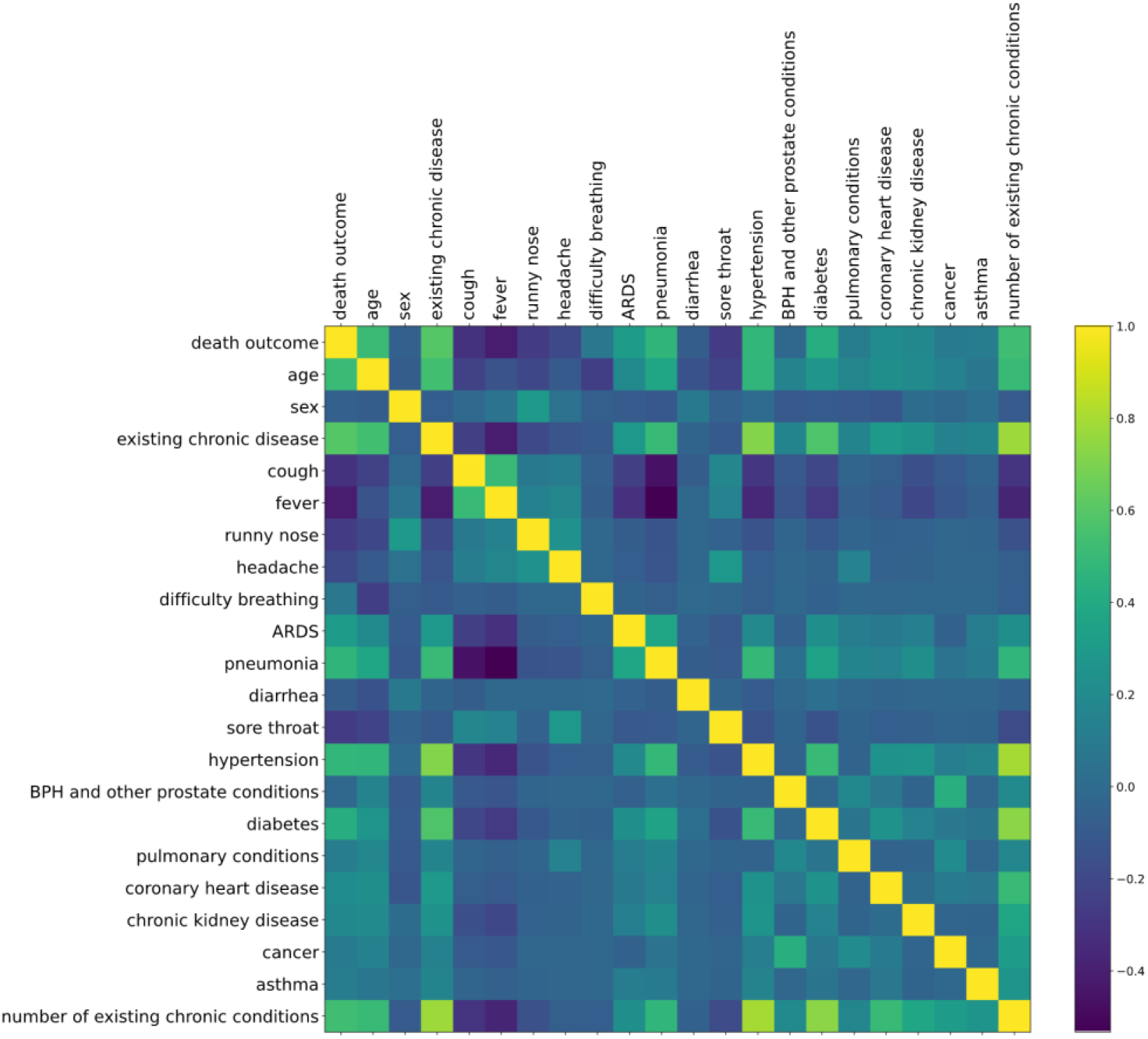
Correlation matrix of patient demographics, symptoms and pre-existing conditions with each other and with an outcome of death.

### 3.2 Prediction Performance on Fully Populated Data

#### 3.2.1 Baseline Characteristics

The fully populated data contains full entries for each category mentioned in the methodology, resulting in a sample of 212 patients. The data is geographically diverse with representation from 17 countries, though 62 patients (29.2%) originate from China alone. The mean (± standard deviation [SD]) age in the sample is 55.9 years (±21.8). The mean age of patients who died of COVID-19 is significantly higher than those who didn’t, at 64.1 (±19.6) against 40.8 (±16.9) respectively. Men comprised 67.9% of the sample. A sizeable 49.5% of the sample suffered from some pre-existing condition, which is overrepresented and 64.6% of patients ultimately died, rendering the final class balance highly skewed.

#### 3.2.2 Performance

The table below (Table 2) outlines the out of sample performance of the two classifiers used in this study on both the full 25 features in the data and a reduced variation with 7 features, using 3-fold cross validation. Random Forest performs better than Logistic Regression across both variations of the data with high accuracy and class specific performance. Both classifiers perform better when reducing the dataset’s dimensionality, more so for Random Forest (92.5% accuracy vs. 89.6%). The best performing model has an AUC of 96.4 and an accuracy of 92.5%, well split between specificity and sensitivity. This dataset however is excessively balanced, so performance is not guaranteed to be representative of live testing.

**Table 2:** Performance of mortality prediction across models measured using 3-fold cross validation on out of sample imbalanced data.

|  | *Full 25 Feature Dataset* |  | *7 Feature Dataset* |  |
| --- | --- | --- | --- | --- |
|  | **Random Forest** | **Logistic Regression** | **Random Forest** | **Logistic Regression** |
| Specificity (%) | 89.3 | 86.7 | 90.7 | 89.3 |
| Sensitivity (%) | 89.8 | 76.6 | 93.4 | 76.6 |
| Accuracy (%) | 89.6 | 80.2 | 92.5 | 81.1 |
| AUC (%) | 96.1 | 89.6 | 96.4 | 89.8 |

### 3.3 Prediction Performance on Large Symptomless Data

As a more representative alternative, the requirement to have symptom data was dropped, and only patients having populated entries for the remaining data categories were kept. This results in greater sample size, but, as will be outlined shortly, lacking data on leading causes of death.

#### 3.3.1 Baseline Characteristics

In total, 5,121 patients are included in the sample. There is representation from 31 countries. The mean (± standard deviation [SD]) age in the sample is 45.6 years (±19.0). The mean age of patients who died of COVID-19 is significantly higher than those who didn’t, at 60.2 (±20.8) against 42.1 (±16.8) respectively. Men comprised 52.8% of the sample. A small 2.4% of the sample suffered from some pre-existing condition, which is underrepresented. Finally, 18.9% of patients ultimately died, which is strongly overrepresented, and in particular is highly overrepresented compared to the proportion of co-morbidities that might help explain it.

#### 3.3.2 Performance

As just mentioned, the proportion of deaths in the sample vastly outweighs the proportion of co-morbidities, and as symptom data is largely missing in this variation of the data, there is a strong lack of explanatory variables. We thus use this sample purely as a large scale, cross-country testing ground for prediction based on age alone. This attribute has already been strongly linked to adverse COVID-19 outcomes, but here it will be specifically benchmarked against the performance of earlier models. The new models tested in this section will form their own custom stratification or linearization of age for prediction purposes, as opposed to semi-arbitrarily drawn age brackets tested for significance in outcome differences, usually found in the literature. The table below (Table 3) outlines model performance on the data. Both featured models perform surprisingly well considering the single variable constraint, with accuracies in the low 70’s and a very favourable breakdown of type 1 and type 2 error. Particularly interesting is their ability to now discern death better than recovery with significantly higher sensitivity than specificity. The data does still feature a high initial proportion of deaths, namely 18.9%, so a live implementation would likely perform worse, though in a hospital setting mortality may remain high. Thus on a large sample of diverse data age is an adequate standalone predictor of mortality, though strongly less effective than a combination model.

**Table 3:**
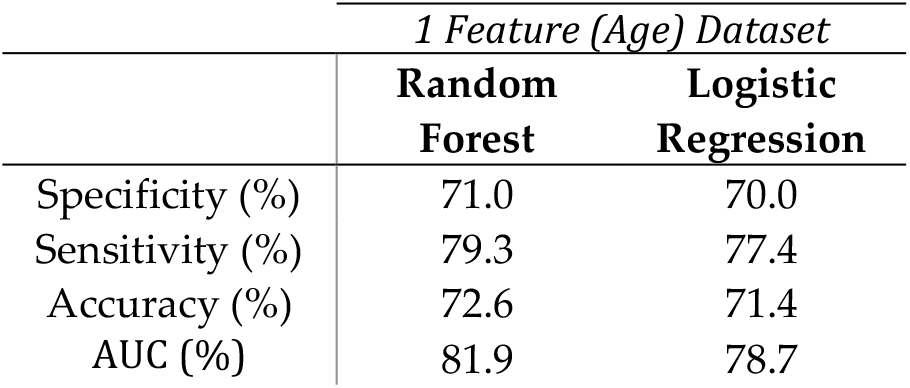
Performance of mortality prediction across models measured using 5-fold cross validation on out of sample imbalanced data.

|  | *1 Feature (Age) Dataset* |  |
| --- | --- | --- |
|  | **Random Forest** | **Logistic Regression** |
| Specificity (%) | 71.0 | 70.0 |
| Sensitivity (%) | 79.3 | 77.4 |
| Accuracy (%) | 72.6 | 71.4 |
| AUC (%) | 81.9 | 78.7 |

## 4 Discussion

Models trained on a high quality 212 patient dataset obtained strong results, with the highest-performing model achieving 93.4% sensitivity and 90.7% specificity on mortality prediction. The dataset however is both small and unrepresentative in its mortality class balance. Performance on a second dataset of large dimensionality, but with slightly overrepresented mortality, was used to gauge how models would perform using only age as a discriminatory characteristic. Surprisingly, age alone acts as a very strong and balanced predictor, with the best model achieving a sensitivity of nearly 80% and an accuracy of nearly 73% on a baseline proportion of mortality of 18.9%. Co-morbidities are highly correlated with age, but in spite of the inevitable omitted variable bias that age would carry in this context, it is noteworthy to find such discriminatory power on the basis of a single attribute. Training the models on age as opposed to developing stratifications for it ex-ante allows an optimal empirical split capable of informing age risk bracketing, and interestingly both Random Forest – which would carry out a more traditional and nonlinear stratification – and Logistic Regression – which would carry out linearization – perform well.

Throughout the study, dimensionality reduction methods greatly improved performance, implying that these methods are imperative for either model discrimination or simply to reduce the number of required variables and patient history to reach meaningful conclusions. This is particularly relevant to models in the literature that make use of laboratory results and other less immediately available data (Knight et. al, 2020).

Across samples, the data employed in this analysis is more diverse than that of many comparable studies, with representation from 17 countries and a comprehensive age range. Unlike existing works (Pourhomayoun & Shakibi, 2020) using the same data, we wish to increase transparency of its flaws. Some primary shortcomings of the dataset are its small size – depending on attribute filtering criteria – the imbalance in classes – with mortality rising to 64.6% of the sample for the highest quality 212 patient dataset – and the prevalence of chronic conditions, which is too high in the 212 patient dataset and too low in the 5,121 patient dataset. Additionally, most data is sourced from the first four months of the pandemic, thus providing no information on later variants of interest, some of which have been linked to greater mortality risk. These limitations are not minute, and caution should be employed in reporting any results based on this widely cited and publicly available dataset. While informative conclusions can be reached, they must be followed by proper disclosure about issues surrounding how representative or small the filtered, high quality data actually is, even if the parent dataset spans millions of entries.

## 5 Conclusion

This study has shown the substantial accuracy machine learning models can bring in the early detection of mortality in COVID-19. The most clinically relevant performance was obtained on a high quality sample of 212 patients with accuracy, sensitivity and specificity all above 90%, though the sample is biased with a roughly even distribution of death and recovery outcomes. As a worst estimate of real life performance, a more representative, imbalanced dataset of 5,121 patients was assembled on the basis solely of age as a mortality predictor, showing that both linear and nonlinear models could reasonably detect death – at 80% sensitivity. The limitations of the data employed in this study have been explored and addressed, providing a cautious warning on future use in the literature.

## Data Availability

Data used for this study is publicly available and described in:
Xu, B., Gutierrez, B., Mekaru, S., Sewalk, K., Goodwin, L., Loskill, A., Cohn, E., Hswen, Y., Hill, S., Cobo, M., Zarebski, A., Li, S., Wu, C., Hulland, E., Morgan, J., Wang, L., O’Brien, K., Scarpino, S., Brownstein, J., Pybus, O., Pigott, D. and Kraemer, M., 2020. Epidemiological data from the COVID-19 outbreak, real-time case information. Scientific Data, 7(1).

https://www.nature.com/articles/s41597-020-0448-0#Sec6

## Notes

### Competing Interest Statement

The authors have declared no competing interest.

### Funding Statement

No external funding was received.

### Author Declarations

The study falls under one of the IRB exempt categories: it is research involving existing data which is publicly available, has been used in other studies published on medRxiv and journals, and has anonymized entries. Indirect identifiers (such as location and demographics) are never mentioned in the study in relation to a single individual or even a group smaller than 200 individuals, so there are also no patient identifiers, direct or indirect, in the body of the submission.

## References

An, C., Lim, H., Kim, D., Chang, J., Choi, Y. and Kim, S., 2020. Machine learning prediction for mortality of patients diagnosed with COVID-19: a nationwide Korean cohort study. Scientific Reports, 10(1).

Barda, N., Riesel, D., Akriv, A., Levy, J., Finkel, U., Yona, G., Greenfeld, D., Sheiba, S., Somer, J., Bachmat, E., Rothblum, G., Shalit, U., Netzer, D., Balicer, R. and Dagan, N., 2020. Developing a COVID-19 mortality risk prediction model when individual-level data are not available. Nature Communications, 11(1).

El-Solh, A., Lawson, Y., Carter, M., El-Solh, D. and Mergenhagen, K., 2020. Comparison of in-hospital mortality risk prediction models from COVID-19. PLOS ONE, 15(12), p.e0244629.

Knight, S., Ho, A., Pius, R., Buchan, I., Carson, G., Drake, T., Dunning, J., Fairfield, C., Gamble, C., Green, C., Gupta, R., Halpin, S., Hardwick, H., Holden, K., Horby, P., Jackson, C., Mclean, K., Merson, L., Nguyen-Van-Tam, J., Norman, L., Noursadeghi, M., Olliaro, P., Pritchard, M., Russell, C., Shaw, C., Sheikh, A., Solomon, T., Sudlow, C., Swann, O., Turtle, L., Openshaw, P., Baillie, J., Semple, M., Docherty, A. and Harrison, E., 2020. Risk stratification of patients admitted to hospital with covid-19 using the ISARIC WHO Clinical Characterisation Protocol: development and validation of the 4C Mortality Score. BMJ, p.m3339.

Pourhomayoun, M. and Shakibi, M., 2021. Predicting mortality risk in patients with COVID-19 using machine learning to help medical decision-making. Smart Health, 20, p.100178.

Xu, B., Gutierrez, B., Mekaru, S., Sewalk, K., Goodwin, L., Loskill, A., Cohn, E., Hswen, Y., Hill, S., Cobo, M., Zarebski, A., Li, S., Wu, C., Hulland, E., Morgan, J., Wang, L., O’Brien, K., Scarpino, S., Brownstein, J., Pybus, O., Pigott, D. and Kraemer, M., 2020. Epidemiological data from the COVID-19 outbreak, real-time case information. Scientific Data, 7(1).

